# Temporal Analysis of COVID-19 Convalescent Plasma Donations Reveals Significant Decrease in Neutralizing Capacity Over Time

**DOI:** 10.1101/2020.10.04.20206011

**Authors:** Roxie C. Girardin, Alan P. Dupuis, Anne F. Payne, Timothy J. Sullivan, Donna Strauss, Monica M. Parker, Kathleen A. McDonough

## Abstract

COVID-19 convalescent plasma (CCP) received approval for use under an Emergency Use Authorization by the FDA for treatment of seriously ill patients. Use of CCP units with a signal-to-cutoff ratio of ≥12 using the Ortho VITROS SARS-CoV-2 IgG test (OVSARS2IgG) is authorized. Little is known about the relationship between this ratio and the neutralizing capacity of plasma/sera against genuine SARS-CoV-2 virus. We measured the neutralizing capacity of 981 samples from 196 CCP donors 7-119 days post initial donation (DPID). Neutralizing capacity was assessed for 50% (PRNT_50_) and 90% (PRNT_90_) reduction of infectious virus using the gold standard plaque reduction neutralization test (PRNT). Importantly, while 32.7%/79.5% (PRNT_90_/PRNT_50_) of donations met the FDA minimum titer of 1:80 initially, only 14.0%/48.8% (PRNT_90_/PRNT_50_) met this cut-off ≥85 DPID. A subset of 91 donations were evaluated using the OVSARS2IgG and compared to PRNT titers for diagnostic accuracy. The correlation of OVSARS2IgG results to neutralizing capacity allowed extrapolation to CCP therapy efficacy results. CCP with OVSARS2IgG ratios in the therapeutically beneficial group had neutralizing titers of ≥1:640 (PRNT_50_) and/or ≥1:80 (PRNT_90_). This information provides a new basis for refining the recommended properties of CCP that is used to treat severe COVID-19.

## Introduction

The emergence of COVID-19 disease, caused by infection with the SARS-CoV-2 virus, has precipitated a global public health crisis. Treatment of COVID-19 primarily consists of supportive care, although several experimental therapies are being tested in clinical trials. Convalescent plasma has long been used as a therapeutic for viral infections when other effective drugs or therapies are absent and has been used to treat SARS infections (1, 2). To date, more than 84,000 patients in the United States have been transfused with COVID-19 convalescent plasma (CCP) (3). The efficacy of CCP has been difficult to establish, but it is thought that CCP has an acceptable safety profile and may provide therapeutic benefit (4-8).

Protective correlates of immunity have not been definitively established for COVID-19, but vaccine trials in non-human primates have shown correlation between neutralizing antibody response and protection from SARS-CoV-2 challenge (9, 10). Thus, the potential therapeutic benefit of CCP is thought to be dependent on the ability of antibodies present in the plasma to neutralize SARS-CoV-2 and block infection. Neutralizing antibodies from recovered COVID-19 patients compete with angiotensin-converting enzyme 2 for binding of receptor binding domain (RBD) on the trimeric spike protein of SARS-CoV-2 (11). Several serological assays have been developed to measure the binding of antibodies in sera/plasma to a variety of SARS-CoV-2 spike antigens, including RBD, the S1 subunit of spike, and trimeric conformations of the spike ectodomain (12, 13). An absolute neutralizing titer associated with therapeutically beneficial CCP has been difficult to elucidate, as many larger published studies characterized none or few of the CCP units for neutralizing titer with live SARS-CoV-2 virus (5, 7, 8). Instead, serological assays that measure the titer of anti-RBD or anti-spike antibodies have served as surrogates, and a lack of widely available standardized reagents and control sera make direct comparison across labs difficult. Several different titers have been recommended after comparison of customized ELISA-based assays to varied microneutralization assays that use SARS-CoV-2 virus, but the relationship between these assays and the neutralizing capacity of sera/plasma as measured by the gold standard plaque reduction neutralization test (PRNT) is not always clear (12-14). This point becomes particularly crucial when selecting CCP with the highest neutralizing capacity and/or avoiding the transfusion of units of CCP with little/no neutralizing capacity (15). Recently, the U.S. Food and Drug Administration (FDA) declared that units of CCP can be qualified as high titer if they have a signal-to-cutoff ratio of ≥12 using the Ortho VITROS SARS-CoV-2 IgG test (OVSARS2IgG) (16). Like other surrogate neutralization assays, the OVSARS2IgG measures antibody binding to the SARS-CoV-2 S1 antigen. A single report has shown concordance between OVSARS2IgG ratios and pseudovirus neutralization, but this test has not been compared to PRNT using genuine SARS-CoV-2 (17).

Symptom severity has been reported to positively correlate with neutralizing capacity convalescent COVID-19 patients, as does male gender and age (18, 19). It has been suggested that these criteria can be used to select the CCP donors most likely to contribute CCP with high neutralizing capacity. However, a specific optimal window post symptom resolution for CCP collection has not been defined (18). There is debate about the stability of neutralizing antibody responses in recovered COVID-19 patients, with conflicting results from different studies. Prior studies lacked serial specimens from the same individual, completed analyses on a small number of serial specimens within a limited time frame, or did not assess neutralizing capacity of paired specimens (14, 19-21). There is great interest in understanding the durability of neutralizing antibody responses in convalescent COVID-19 patients, as there is concern that rapid decay in neutralizing capacity could result in susceptibility to re-infection with SARS-CoV-2. Additionally, significant decreases in the neutralizing antibody capacity of convalescent COVID-19 patients would render time post symptom resolution a critical factor in CCP donor selection.

The goals of this study are to characterize the neutralizing capacity of serial donations of CCP using PRNT with genuine SARS-CoV-2, to evaluate the CCP-qualifying criteria of the OVSARS2IgG test at different levels of neutralizing capacity, and to correlate PRNT titer and OVSARS2IgG ratios to data on the therapeutic efficacy of CCP compiled by the Mayo Clinic/US EAP COVID-19 Plasma Consortium (US-EAP-CPC).

## Results and Discussion

### The durability of the neutralizing capacity of CCP donors

Of the 981 specimens analyzed ranging up to 119 DPID, 61.1% were contributed by males, 38.4% were contributed by females, and 0.5% were contributed by individuals that did not specify gender. Specimens contributed by males had significantly greater mean neutralizing capacities (P value <0.0001) than those contributed by females. The mean age of the study population was 48 years. Age was weakly, positively correlated with neutralizing titer (PRNT_50_ Spearman r 0.2129, P value <0.0001; PRNT_90_ Spearman r 0.1988, P value <0.0001). Neutralizing antibodies were not detected in 1.3% of CCP tested at the minimum dilution (1:20) screened in this study. As a population, 25.8% (PRNT_90_) to 73.0% (PRNT_50_) of donations had a PRNT titer of ≥1:80. Comparatively, 9.5% (PRNT_90_) to 51.4% (PRNT_50_) of all donations had a PRNT titer of ≥1:160.

Neutralizing capacities of donations were compared in two-week intervals at the PRNT_50_ (Table 1) and PRNT_90_ levels (Supplemental Table 1). The proportion of specimens that meet the FDA minimal 1:80 and recommended 1:160 cut-offs decreased over time at both levels of neutralization. A significant proportion (23.4%) of donations decreased ≥4-fold in PRNT_50_ titer, while fewer (8.5%) had a decreased PRNT_90_ titer versus their initial draw (Supplemental Table 2 and 3). Mean PRNT_50_ neutralizing titers decreased significantly over time (P <0.0001) (Figure 1A and B). The most significant decreases in PRNT_50_ titer occurred at ≥43 DPID, suggesting a 6-week window, likely corresponding to 3 -9 weeks post symptom onset, is optimal for maximizing collection of high titer CCP. While the decrease in PRNT_90_ neutralizing titers was not statistically significant (P= 0.0661) (Supplemental Figure 1A and B), all donors with titers ≥1:320 experienced decreases in PRNT_90_ titer by their final draw (Supplemental Figure 1B). While 9.2% (18/196) of CCP donors converted from positive to negative for neutralizing activity at the PRNT_90_ level, none of the donors converted from positive to negative for neutralizing activity at the PRNT_50_ level at any time point. Loss of neutralizing capacity at PRNT_90_ level occurred at ≥61 DPID for 15 of 18 donors. While low a PRNT_50_ titer may render a unit of CCP undesirable for use as a COVID-19 therapeutic, these results suggest that individuals that produce neutralizing antibody retain some SARS-CoV-2 neutralizing capacity for up to 119 DPID.

**Table 1:** The distribution of PRNT_50_ titers of CCP donations over time.

| PRNT<sub>50</sub> | 0 | 1-14d | 15-28d | 29-42d | 43-56d | 57-70d | 71-84d | >=85d |
| --- | --- | --- | --- | --- | --- | --- | --- | --- |
| <20 | 2.1% (4) | 1.4% (3) | 2.8% (4) | 1.8% (2) | 0% (0) | 0% (0) | 0% (0) | 0% (0) |
| 20 | 3.1% (6) | 6.0% (13) | 5.5% (8) | 9.0% (10) | 8.0% (9) | 12.2% (10) | 16.7% (12) | 11.6% (5) |
| 40 | 15.4% (30) | 16.5% (36) | 15.2% (22) | 16.2% (18) | 23.2% (26) | 17.1% (14) | 20.8% (15) | 39.5% (17) |
| 80 | 15.9% (31) | 17.4% (38) | 24.8% (36) | 23.4% (26) | 23.2% (26) | 35.4% (29) | 25.0% (18) | 16.3% (7) |
| 160 | 17.4% (34) | 22.0% (48) | 15.9% (23) | 21.6% (24) | 24.1% (27) | 13.4% (11) | 18.1% (13) | 16.3% (7) |
| 320 | 12.3% (24) | 12.8% (28) | 11.7% (17) | 12.6% (14) | 11.6% (13) | 8.5% (7) | 6.9% (5) | 11.6% (5) |
| 640 | 7.7% (15) | 4.6% (10) | 5.5% (8) | 6.3% (7) | 4.5% (5) | 11.0% (9) | 9.7% (7) | 2.3% (1) |
| >640 | 26.2% (51) | 19.3% (42) | 18.6% (27) | 9.0% (10) | 5.4% (6) | 2.4% (2) | 2.8% (2) | 2.3% (1) |
| ≥80 | 79.5% (155) | 76.1% (166) | 76.6% (111) | 73.0% (81) | 68.8% (77) | 70.7% (58) | 62.5% (45) | 48.8% (21) |
| ≥160 | 63.6% (124) | 58.7% (128) | 51.7% (75) | 49.5% (55) | 45.5% (51) | 35.4% (29) | 37.5% (27) | 32.6% (14) |

**Table 2:** The diagnostic accuracy profiles of the OVSARS2IgG test at two cut-offs versus the gold standard PRNT_50_ titer. 95% confidence intervals are reported for all comparisons of diagnostic accuracy.

| PRNT<sub>50</sub> | NT ≥20,<br>OVSARS2Ig<br>G ratio ≥12 | NT ≥20,<br>OVSARS2Ig<br>G ratio >18.45 | NT ≥80,<br>OVSARS2Ig<br>G ratio ≥12 | NT ≥80,<br>OVSARS2Ig<br>G ratio >18.45 | NT ≥160,<br>OVSARS2Ig<br>G ratio ≥12 | NT ≥160,<br>OVSARS2Ig<br>G ratio >18.45 |
| --- | --- | --- | --- | --- | --- | --- |
| Sensitivity | 65.5%<br>(0.5483-0.7476) | 48.8%<br>(0.3841-0.5931) | 85.5%<br>(0.7466-0.9217) | 66.1%<br>(0.5372-0.7666) | 89.8%<br>(0.7824-0.9556) | 77.6%<br>(0.6412-0.8698) |
| Specificity | 100.0%<br>(0.6457-1.000) | 100.0%<br>(0.6457-1.000) | 93.1%<br>(0.7804-0.9877) | 100.0%<br>(0.8830-1.000) | 73.8%<br>(0.5893-0.8470) | 92.9%<br>(0.8099-0.9574) |
| Positive Predictive<br>Value | 100.0%<br>(0.9347-1.000) | 100.0%<br>(0.9143-1.000) | 96.4%<br>(0.8768-0.9935) | 100.0%<br>(0.9143-1.000) | 80.0%<br>(0.6764-0.8845) | 92.7%<br>(0.8057-0.9748) |
| Negative Predictive<br>Value | 19.4%<br>(0.09753-0.3503) | 14.0%<br>(0.06951-0.2619) | 75.0%<br>(0.5893-0.8625) | 58.0%<br>(0.4423-0.7062) | 86.1%<br>(0.7134-0.9392) | 78.0%<br>(0.6476-0.8725) |
| P value (Fisher's<br>Exact Test) | 0.0010 | 0.0151 | <0.0001 | <0.0001 | <0.0001 | <0.0001 |

**Figure 1:**
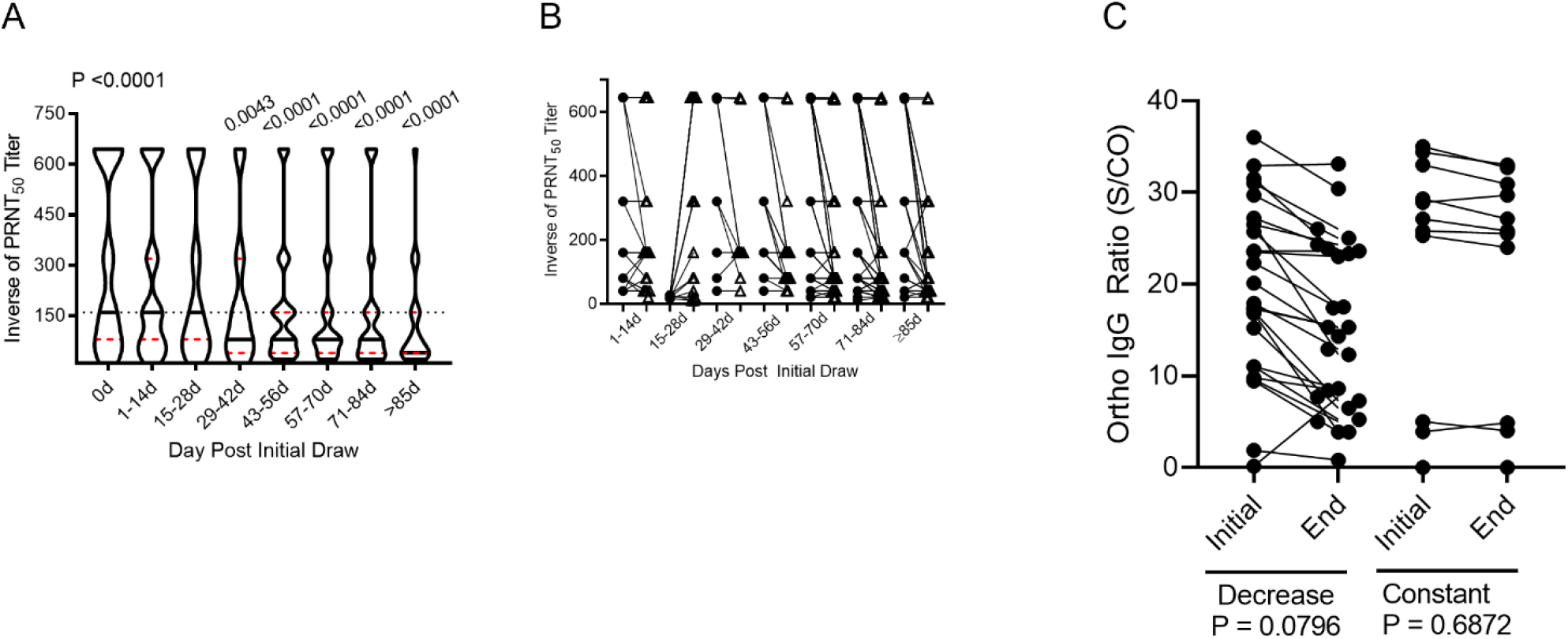
Neutralizing capacity of CCP donations decreases significantly over time. A. Distribution of PRNT_50_ titers in two-week intervals (d, days) of CCP donations. The Kruskal-Wallis test with Dunn’s correction for multiple comparisons was applied. Each of the two-week time periods was compared to initial collection (0d) and statistically significant differences are noted by the P value above the group. B. Initial (circles) and final (triangles) PRNT_50_ titers for all donors. C. A subset of study participants were assessed using the OVSARS2IgG test and are grouped by specimen pairs that had decreases in PRNT titers (Decrease) or those that were constant (Constant). The Mann-Whitney test was applied; P values are indicated. A-B: n=91. C n=91.

### Ortho VITROS SARS-CoV-2 IgG test and neutralizing capacity of sera

A subset of specimens was evaluated using the FDA-approved OVSARS2IgG test at FDA and Mayo Clinic/US-EAP-CPC derived cut-offs. The OVSARS2IgG ratios of sera from initial draws were compared to the OVSARS2IgG ratios of a subsequent draw where neutralizing titer remained constant or decreased. While most sera pairs with decreases in neutralizing capacity also had decreases in OVSARS2IgG ratios, the difference was not statistically significant (Figure 1C). Sera with less than FDA mandated OVSARS2IgG ratio (<12) had significantly lower PRNT_50_ and PRNT_90_ titers than sera with a ratio ≥12 (Figure 2A and D). Next, the distribution of PRNT titers was analyzed according to the OVSARS2IgG ratio ranges used by the Mayo Clinic/US-EAP-CPC in a recent report on the efficacy of CCP (5). The PRNT_50_ and PRNT_90_ titers of sera with an OVSARS2IgG ratio >18.45 were significantly higher than those with ratios <4.62 or 4.62-18.45 (Figure 2B and E). The samples analyzed include an approximate representation of the distribution of neutralizing titers of the larger study population, but performance characteristics could be altered if a greater proportion of specimens with very high or very low neutralizing titers were assayed instead.

**Figure 2:**
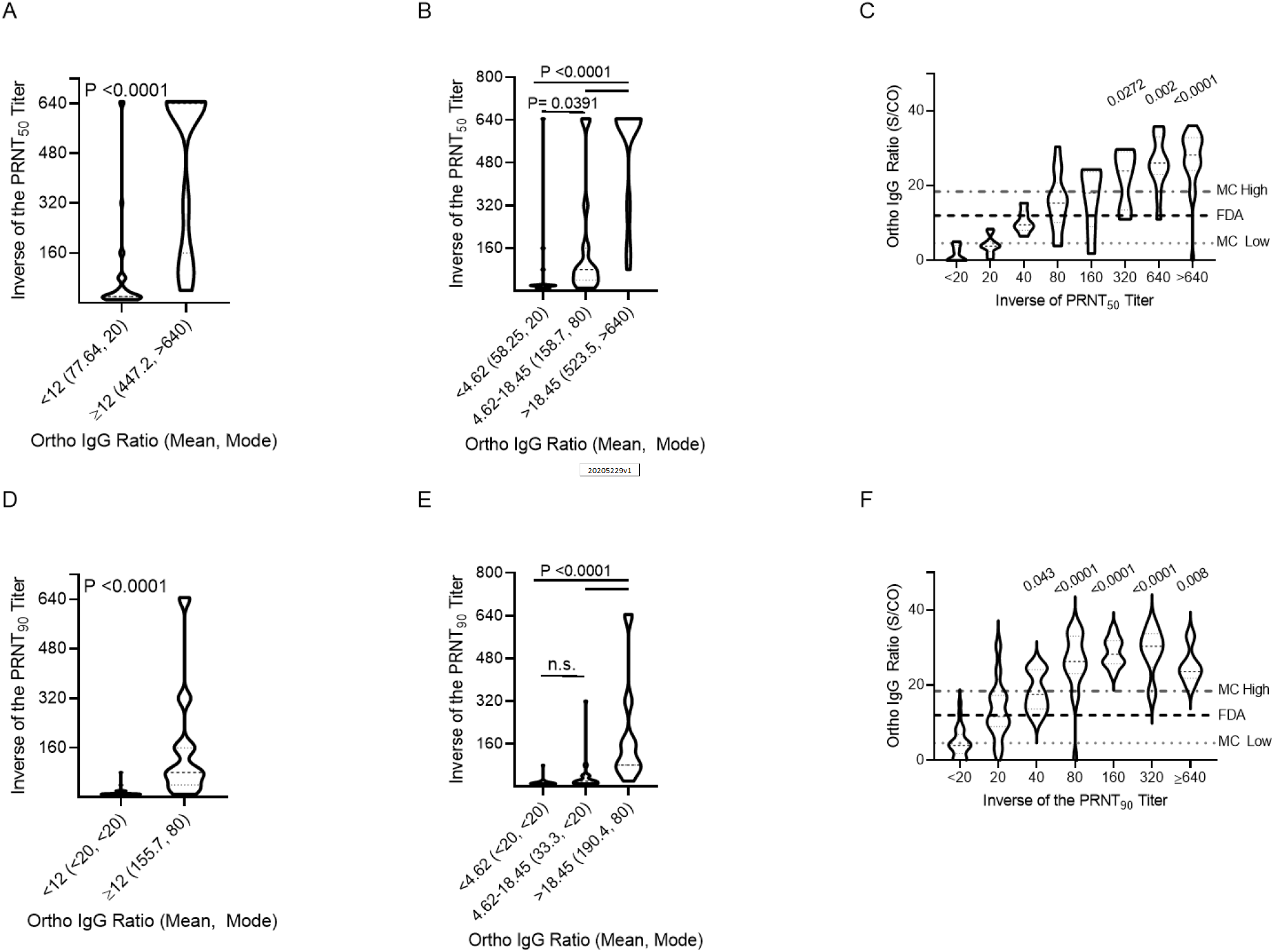
Comparison of the OVSARS2IgG test to the neutralizing capacity of CCP. A. Distribution of PRNT_50_ titers in groups with OVSARS2IgG ratios using the FDA cut-off. The Mann-Whitney test was applied; P values are indicated. B. Distribution of PRNT_50_ titers in groups with OVSARS2IgG ratios defined by the Mayo Clinic/US-EAP-CPC. The Kruskal-Wallis test with Dunn’s correction for multiple comparisons was applied; P values are indicated. C. Distribution of PRNT_50_ titers across all OVSARS2IgG test ratios. The Kruskal-Wallis test with Dunn’s correction for multiple comparisons was usedd to compare PRNT_50_ ≥20 to groups with PRNT_50_ <20. P values are indicated. D. As in A, but for PRNT_90_ titers. E. As in B, but for PRNT_90_ titers. F. As in C, but for PRNT_90_ titers. n=91.

The accuracy of the OVSARS2IgG test to qualify high titer CCP was compared to the reference standard PRNT at both levels of neutralizing capacity. The FDA-established OVSARS2IgG ratio of ≥12 and the Mayo Clinic/US-EAP-CPC derived ratio of >18.45 were used in the performance assessment. The ability of the OVSARS2IgG test to correctly identify sera with any neutralizing capacity (inverse of the neutralizing titer (NT) NT≥20), or those with titers at the FDA recommended levels (NT≥80 or NT≥160) at the PRNT_50_ (Table 2) and PRNT_90_ levels was determined (Supplemental Table 4). The FDA cut-off of 12 resulted in 100% specificity and positive predictive value for NT≥20 and NT≥80 at the PRNT_50_ level. Sensitivity and negative predictive value improved when capturing specimens with NT≥80 or NT≥160 compared to NT≥20 at the PRNT_50_ level.

CCP with an OVSARS2IgG ratio >18.45 significantly correlates with improved outcomes in patients transfused shortly after hospitalization (5). While our results support the use of an OVSARS2IgG ratio ≥12 to exclude CCP with no neutralizing capacity from therapeutic use, we determined whether an OVSARS2IgG ratio >18.45 provided additional discriminatory power. Significantly, the >18.45 cut-off had improved specificity and positive predictive value for specimens with NT≥160 (Table 2). It also had better specificity and positive predictive value than a cut-off of ≥12 for specimens with a NT≥20 or NT≥80 at the PRNT_90_ level (Supplemental Table 4). The mean PRNT_50_ titer of the >18.45 group is 1:523.5 (mode >1:640), while the mean PRNT_90_ titer is 1:190.4 (mode 1:80) (Figure 2B and E). Analysis of the distribution of neutralizing titers compared to the OVSARS2IgG ratios and the ranges associated with efficacy by the Mayo Clinic/US-EAP-CPC revealed that sera with the highest neutralizing capacity have OVSARS2IgG ratios that are significantly higher than sera with low neutralizing capacity (Figure 2 C and F). These direct comparisons between the neutralizing capacity of sera, their corresponding OVSARS2IgG ratios, and the patient outcome data released by the Mayo Clinic/US-EAP-CPC support updating recommendations for CCP use to specify that a PRNT_90_ titer ≥1:80 and/or a PRNT_50_ titer ≥1:640 qualify as high titer CCP.

The analysis presented herein clarifies the functional relationships between live SARS-CoV-2 virus neutralizing titers and the only current FDA approved surrogate neutralization test for qualifying CCP. While an OVSARS2IgG test ratio ≥12 excluded all tested specimens with a NT<20 at the PRNT_50_ level, this ratio does not exclude specimens with low neutralizing capacity and uncertain therapeutic efficacy from being labeled high titer CCP. Rather, high titer CCP donations are better characterized by an OVSARS2IgG test ratio >18.45, which maximizes specificity and positive predictive value at both levels of neutralizing capacity measured. This recommendation is supported by our observation that CCP in a therapeutically beneficial treatment group is most likely to have PRNT_90_ titer ≥1:80 and/or a PRNT_50_ titer ≥1:640. Along with previously established guidelines for donor selection (18), consideration of time since disease resolution will further refine donor selection to yield high titer CCP, particularly when CCP donations cannot be tested for their neutralizing capacity using live SARS-CoV-2.

## Methods

### Study Specimens

Specimens were collected from individuals who met all FDA donor eligibility requirements (21CFR 630.10 and 21 CFR 630.15) and qualifications in accordance with the guidelines of the New York Blood Center (NYBC). Donors were required to present documentation of a positive SARS-CoV-2 PCR diagnostic test or positive serologic test after recovery and to have been symptom free for at least two weeks. Donors contributed CCP/sera at will, with a minimum of 7 days between serial contributions. Specimens were stored at -20°C until tested. Testing at the Wadsworth Center was done under protocol 20-021 with approval from the NYSDOH Institutional Review Board.

### PRNT Analysis

Participants were selected at random from a de-identified list of residual clinical specimens submitted for anti-SARS-CoV-2 antibody testing at the Wadsworth Center Laboratory. Groups of donors with varying intervals between initial donation and final donation were retrospectively selected for study: 14-35d (n=45), 36-60d (n=27), 61-75d (n=55), and >75 (n=69). Sample selection and testing was blinded to clinical data including age, gender, and SARS-CoV-2 antibody test results from the Wadsworth Center clinical assay.

PRNT analysis was conducted by mixing 100 ul of 200 PFUs of SARS-CoV-2, isolate USA-WA1/2020 (BEI Resources, NR181 52281) with 100 ul of 2-fold serially diluted test sera and incubated at 37°C in 5% CO2 for one hour. Confluent Vero E6 cells (C1008, ATCC CRL-1586) seeded in 6 well plates were inoculated with 100 ul of the virus:serum mixture and adsorption proceeded for one hour at 37°C in 5% CO2. A 0.6% agar overlay prepared in maintenance medium (Eagle’s Minimal Essential Medium, 2% heat-inactivated FBS, 100 µg/ml Penicillin G, 100 U/ml Streptomycin) was added after adsorption and the assay was incubated at 37°C in 5% CO2. A second agar overlay with 0.2% Neutral red added was added two days post infection. After an additional day of incubation, the number of plaques in each well were recorded. The titer was reported as the inverse of the highest dilutions of sera providing 50% (PRNT_50_) or 90% (PRNT_90_) viral plaque reduction relative to virus-only infection. Normal human serum was used as a negative control and previously characterized COVID-19 patient sera was used as a positive control in each assay.

### Ortho VITROS SARS-CoV-2 IgG testing

A subset of ∼10% of donor sera was selected for analysis using the Ortho VITROS SARS-CoV-2 IgG test (OVSARS2IgG). Specimens with missing/indeterminate PRNT titers were excluded from analysis. The subset was selected so it approximated the total study population with regards to neutralization at/above the FDA minimal 1:80 titer at the PRNT_50_ level (total study population = 73%, subset = 68.1%). Additionally, the subset included 11 matched pairs with steady PRNT titers and 21 matched pairs with decreases in PRNT titers (late versus early donation). Two selected donations had insufficient volume for testing, so were replaced with specimens with equivalent titers selected at random from NYBC specimens subjected to PRNT. DPID ranged from 0-114d for the subset. The OVSARS2IgG test was run on the Ortho VITROS 5600 instrument according to manufacturer’s instructions and the signal-to-cutoff ratio was automatically calculated by the system. The laboratorians completing the OVSARS2IgG were blinded to clinical information on the specimens, including age, gender, SARS-CoV-2 antibody test results from the Wadsworth Center clinical assay, PRNT results, and date of collection.

### Statistical Analyses

Correlation between age and neutralizing titer was assessed with a two-tailed Spearman’s r test. Statistical significance of correlations and comparisons between PRNT titers, DPID, and OVSARS2IgG ratios were computed using the Kruskal-Wallis test with Dunn’s correction for multiple comparisons when comparing three or more groups. The Mann-Whitney test was applied when comparing two groups of a continuous variable, and the Kolmogorov-Smirov test was used when comparing two groups of a discrete variable. The sensitivity, specificity, positive predictive value, and negative predictive value were calculated using the Wilson-Brown method for calculating 95% confidence intervals. A two-sided Fisher’s exact test was used to assess significance of the effect sizes calculated.

## Data Availability

Yes

## Author Contributions

Study design: RCG. Experimentation: RCG, ADP, AFP, TJS. Materials/support: M.P., D.S., KAM. Data analysis: RCG. All authors contributed to the writing and/or editing of the manuscript.

## Acknowledgements

We thank the Tissue Culture and Media Core of the Wadsworth Center for the provision of cultured cells and reagents, Jess Machowski, Sean Bialosuknia, and the Diagnostic Immunology team at Wadsworth Center for continued support, the Arbovirus and Rabies Laboratories at Wadsworth for support and technical assistance, and the many selfless plasma donors who have come forward in support of their communities in the fight against COVID-19. Support was provided by CDC ELC CARES for COVID-19.

## Notes

### Competing Interest Statement

The authors have declared no competing interest.

### Author Declarations

Testing at the Wadsworth Center was done under protocol 20-021 with approval from the NYSDOH Institutional Review Board.

## References

1. Cheng Y, Wong R, Soo YO, Wong WS, Lee CK, Ng MH, et al. Use of convalescent plasma therapy in SARS patients in Hong Kong. Eur J Clin Microbiol Infect Dis. 2005;24(1):44–6.

2. Yeh KM, Chiueh TS, Siu LK, Lin JC, Chan PK, Peng MY, et al. Experience of using convalescent plasma for severe acute respiratory syndrome among healthcare workers in a Taiwan hospital. J Antimicrob Chemother. 2005;56(5):919–22.

3. Clinic M. https://www.uscovidplasma.org/,. Accessed Septermber, 2020.

4. Duan K, Liu B, Li C, Zhang H, Yu T, Qu J, et al. Effectiveness of convalescent plasma therapy in severe COVID-19 patients. Proceedings of the National Academy of Sciences. 2020;117(17):9490–6.

5. Joyner MJ, Bruno KA, Klassen SA, Kunze KL, Johnson PW, Lesser ER, et al. Safety Update: COVID-19 Convalescent Plasma in 20,000 Hospitalized Patients. Mayo Clinic Proceedings. 2020;95(9):1888–97.

6. Joyner MJ, Senefeld JW, Klassen SA, Mills JR, Johnson PW, Theel ES, et al. Effect of Convalescent Plasma on Mortality among Hospitalized Patients with COVID-19: Initial Three-Month Experience. medRxiv. 2020.

7. Maor Y, Cohen D, Paran N, Israely T, Ezra V, Axelrod O, et al. Compassionate use of convalescent plasma for treatment of moderate and severe pneumonia in COVID-19 patients and association with IgG antibody levels in donated plasma. EClinicalMedicine. 2020:100525.

8. Salazar E, Christensen PA, Graviss EA, Nguyen DT, Castillo B, Chen J, et al. Treatment of Coronavirus Disease 2019 Patients with Convalescent Plasma Reveals a Signal of Significantly Decreased Mortality. Am J Pathol. 2020.

9. Gao Q, Bao L, Mao H, Wang L, Xu K, Yang M, et al. Rapid development of an inactivated vaccine for SARS-CoV-2. bioRxiv. 2020:2020.04.17.046375.

10. Mercado NB, Zahn R, Wegmann F, Loos C, Chandrashekar A, Yu J, et al. Single-shot Ad26 vaccine protects against SARS-CoV-2 in rhesus macaques. Nature. 2020.

11. Ju B, Zhang Q, Ge J, Wang R, Sun J, Ge X, et al. Human neutralizing antibodies elicited by SARS-CoV-2 infection. Nature. 2020;584(7819):115–9.

12. Amanat F, Stadlbauer D, Strohmeier S, Nguyen T, Chromikova V, McMahon M, et al. A serological assay to detect SARS-CoV-2 seroconversion in humans. medRxiv. 2020:2020.03.17.20037713.

13. Salazar E, Kuchipudi SV, Christensen PA, Eagar T, Yi X, Zhao P, et al. Convalescent plasma anti-SARS-CoV-2 spike protein ectodomain and receptor binding domain IgG correlate with virus neutralization. J Clin Invest. 2020.

14. Lee WT, Girardin RC, Dupuis AP, Kulas KE, Payne AF, Wong SJ, et al. Neutralizing Antibody Responses in COVID-19 Convalescent Sera. medRxiv. 2020:2020.07.10.20150557.

15. Liu STH, Lin HM, Baine I, Wajnberg A, Gumprecht JP, Rahman F, et al. Convalescent plasma treatment of severe COVID-19: a propensity score-matched control study. Nat Med. 2020.

16. Administration USDoHaHSFaD. EUA 26382: Emergency Use Authorization (EUA) Request (original request 8/12/20; amended request 8/23/20)

17. Luchsinger LL, Ransegnola B, Jin D, Muecksch F, Weisblum Y, Bao W, et al. Serological Assays Estimate Highly Variable SARS-CoV-2 Neutralizing Antibody Activity in Recovered COVID19 Patients. J Clin Microbiol. 2020.

18. Klein SL, Pekosz A, Park HS, Ursin RL, Shapiro JR, Benner SE, et al. Sex, age, and hospitalization drive antibody responses in a COVID-19 convalescent plasma donor population. J Clin Invest. 2020.

19. Wang X, Guo X, Xin Q, Pan Y, Hu Y, Li J, et al. Neutralizing Antibodies Responses to SARS-CoV-2 in COVID-19 Inpatients and Convalescent Patients. Clin Infect Dis. 2020.

20. Wajnberg A, Amanat F, Firpo A, Altman D, Bailey M, Mansour M, et al. SARS-CoV-2 infection induces robust, neutralizing antibody responses that are stable for at least three months. medRxiv. 2020:2020.07.14.20151126.

21. Wu F, Wang A, Liu M, Wang Q, Chen J, Xia S, et al. Neutralizing antibody responses to SARS-CoV-2 in a COVID-19 recovered patient cohort and their implications. medRxiv. 2020:2020.03.30.20047365.

