## Supplemental Data for "Temporal Analysis of COVID-19 Convalescent Plasma Donations Reveals Significant Decrease in Neutralizing Capacity Over Time"

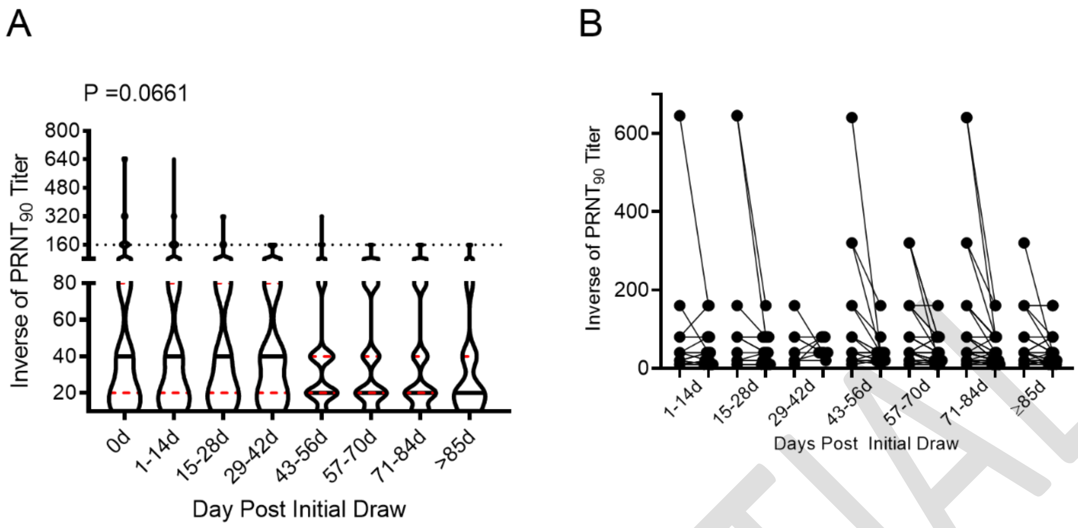


**Supplemental Figure 1: The neutralizing capacity of CCP donations decreases significantly over time.** A. The distribution of $PRNT_{90}$ titers in two-week intervals (d, days) of CCP donations is displayed. The Kruskal-Wallis test with Dunn's correction for multiple comparisons was applied to measure statistical significance. Each of the two-week time periods was compared to initial collection (0d) and statistically significant differences are noted by the P value above the group. B. Initial and end titers for all CCP donors are displayed for $PRNT_{90}$.

CONFIDENTIAL

| PRNT$_{90}$ | 0 | 1-14d | 15-28d | 29-42d | 43-56d | 57-70d | 71-84d | >=85d |
|---|---|---|---|---|---|---|---|---|
| <20 | 19.4% (38) | 19.6% (43) | 15.1% (22) | 15.3% (17) | 12.5% (14) | 18.3% (15) | 22.2% (16) | 25.6% (11) |
| 20 | 27.6% (54) | 30.1% (66) | 34.2% (50) | 28.8% (32) | 42.0% (47) | 39.0% (32) | 38.9% (28) | 39.5% (17) |
| 40 | 20.4% (40) | 22.4% (49) | 21.2% (31) | 28.8% (32) | 31.3% (35) | 20.7% (17) | 18.15 (13) | 20.9% (9) |
| 80 | 15.8% (31) | 16.0% (35) | 21.9% (32) | 19.8% (22) | 8.9% (10) | 18.3% (15) | 15.3% (11) | 9.3 % (4) |
| 160 | 9.7% (19) | 8.7% (19) | 4.8% (7) | 7.2% (8) | 4.5% (5) | 3.7% (3) | 5.6% (4) | 4.7% (2) |
| 320 | 4.1% (8) | 2.7% (6) | 2.7% (4) | 0.0% (0) | 0.9% (1) | 0.0% (0) | 0.0% (0) | 0.0% (0) |
| 640 | 1.5% 3) | 0.5% (1) | 0.0% (0) | 0.0% (0) | 0.0% (0) | 0.0% (0) | 0.0% (0) | 0.0% (0) |
| >640 | 1.5% (3) | 0.0% (0) | 0.0% (0) | 0.0% (0) | 0.0% (0) | 0.0% (0) | 0.0% (0) | 0.0% (0) |
| ≥80 | 32.7% (64) | 27.9% (61) | 29.5% (43) | 27.0% (30) | 14.3% (16) | 22.0% (18) | 20.8% (15) | 14.0% (6) |
| ≥160 | 16.8% (33) | 11.9% (26) | 7.5% (11) | 7.2% (8) | 5.4% (6) | 3.7% (3) | 5.6% (4) | 4.7% (2) |

**Supplemental Table 1: The distribution of PRNT$_{50}$ titers of CCP donations over time.**

CONFIDENTIAL

| $PRNT_{50}$ | 1-14d | 15-28d | 29-42d | 43-56d | 57-70d | 71-84d | >=85d |
|---|---|---|---|---|---|---|---|
| No Change | 87.2% (190) | 81.4% (118) | 73.0% (81) | 67.9% (76) | 67.1% (55) | 66.7% (48) | 53.5% (23) |
| ≥4-Fold Decrease | 10.1% (22) | 17.2% (25) | 27.0% (30) | 32.1% (36) | 31.7% (26) | 33.3% (24) | 46.5% (20) |
| ≥2-Fold Increase | 2.8% (6) | 1.4% (2) | 0% (0) | 0% (0) | 1.2% (1) | 0% (0) | 0% (0) |

**Supplemental Table 2: CCP donations with a ≥4-fold decrease in $PRNT_{50}$ titer presented in two-week intervals.**

CONFIDENTIAL

| $PRNT_{90}$ | 1-14d | 15-28d | 29-42d | 43-56d | 57-70d | 71-84d | >=85d |
|---|---|---|---|---|---|---|---|
| No Change | 95.4% (204) | 92.5% (135) | 90.1% (100) | 81.3% (91) | 89.0% (73) | 90.3% (65) | 93.0% (40) |
| ≥4-Fold Decrease | 3.7% (8) | 7.5% (11) | 9.0% (10) | 17.9% (20) | 9.8% (8) | 9.7% (7) | 7.0% (3) |
| ≥2-Fold Increase | 0.9% (2) | 0% (0) | 0.9% (1) | 0.9% (1) | 1.2% (1) | 0% (0) | 0% (0) |

**Supplemental Table 3: CCP donations with a ≥4-fold decrease in $PRNT_{90}$ titer presented in two-week intervals.**

CONFIDENTIAL

| $PRNT_{90}$ | NT ≥20, OVSARS2IgG ratio ≥12 | NT ≥20, OVSARS2IgG ratio >18.45 | NT ≥80, OVSARS2IgG ratio ≥12 | NT ≥80, OVSARS2IgG ratio >18.45 | NT ≥160, OVSARS2IgG ratio ≥12 | NT ≥160, OVSARS2IgG ratio >18.45 |
|---|---|---|---|---|---|---|
| Sensitivity | 85.5% (0.7466-0.9217) | 91.1% (0.7927-0.9649) | 97.4% (0.8682-0.9987) | 89.7% (0.7642-0.9594) | 100.0% (0.8454-1.000) | 95.2% (0.7733-0.9976) |
| Specificity | 93.1% (0.7804-0.9877) | 100.0% (0.9229-1.000) | 67.3% (0.5376-0.7848) | 88.5% (0.7703-0.9460) | 52.9% (0.4124-0.6433) | 70.0% (0.5846-0.7946) |
| Positive Predictive Value | 96.4% (0.8768-0.9935) | 100.0% (0.9143-1.000) | 69.1% (0.5597-0.7972) | 85.4% (0.7156-0.9312) | 39.6% (0.2759-0.5306) | 48.8% (0.3425-0.6352) |
| Negative Predictive Value | 75.0% (0.5893-0.8625) | 92.0% (0.8116-0.9685) | 97.2% (0.8583-0.9986) | 92.0% (0.8116-0.9685) | 100.0% (0.9036-1.000) | 98.0% (0.8950-0.9990) |
| P value (Fisher's Exact Test) | <0.0001 | <0.0001 | <0.0001 | <0.0001 | <0.0001 | <0.0001 |

**Supplemental Table 4: The diagnostic accuracy profiles of the OVSARS2IgG test at two cut-offs versus the gold standard $PRNT_{90}$ titer.**

CONFIDENTIAL
